# Comparative Analysis of Decision Trees on Two COVID-19 Symptom Datasets

**DOI:** 10.1101/2023.06.02.23290867

**Authors:** Somchai Saengamnatdej, Phuangphet Waree Molee, Prateep Warnnissorn

## Abstract

**Objective:** This study compares decision trees on two COVID-19 symptom datasets to assess their performance and feature importance in predicting and understanding infection patterns.

**Methods:** We created decision trees on Israeli and Swedish COVID-19 infection datasets. Performance metrics were used to assess their predictive capabilities, and feature importance analysis identified significant variables in the decision-making process.

**Results:** The study observed different performance levels of decision trees on the COVID-19 datasets. The Swedish dataset achieved high accuracy and F1-score without hyperparameter tuning, while the Israeli dataset improved significantly with Extreme Gradient Boosting. Dataset characteristics impact the selection of an optimal decision tree algorithm. The key variable in both datasets was sore throat.

**Conclusion:** This study compares decision trees on COVID-19 infection datasets, emphasizing the importance of dataset characteristics in selecting an optimal algorithm. Identifying significant features enhances understanding of infection patterns, benefiting decision-making and prediction accuracy in infectious disease analysis.

## Introduction

Infectious diseases pose significant challenges to public health systems worldwide, leading to substantial morbidity and mortality rates. The accurate prediction and understanding of infection patterns are crucial for effective disease management, prevention, and control strategies. Machine learning algorithms, such as decision trees, have emerged as valuable tools for analyzing complex datasets and making predictions in the field of infectious diseases.

Since the beginning of the pandemic, COVID symptoms have evolved as a result of virus evolution, vaccination, and immunity from earlier infections. The virus evolved, as evidenced by the symptoms shifting from the lower respiratory tract in the early virus strain to the upper airways in later virus strains. The severity of the lungs is also reduced by vaccination an d protection from prior infections, leaving only milder upper respiratory symptoms like fever, myalgia, tiredness, sneezing, sore throat, and cough. These symptoms may be a symptom of various viral diseases and are not unique to COVID-19 (1).

Decision trees are widely used machine learning algorithms that facilitate interpretable and intuitive decision-making processes. These algorithms recursively split the dataset based on the values of specific features, creating a hierarchical structure of decision rules. By capturing the relationships between input variables and infection outcomes, decision trees provide valuable insights into the underlying patterns and mechanisms of infections.

The decision tree offers several key advantages. Firstly, it excels in simplifying complex relationships between multiple variables. Additionally, decision trees are known for their ease of understanding and interpretation. They also possess effective data handling and processing capabilities (2). However, one notable limitation of decision trees is their binary split nature, meaning only one model can be assigned for making predictions at any given point (3). While it is possible to implement multiway splits, they often result in smaller data fragments at each subsequent level and would potentially lead to insufficient data for the next level (4). Despite this limitation, decision trees remain a valuable tool in many scenarios.

High-accuracy machine learning-based diagnostic predictions for COVID-19 have been reported in the literature (5). One study analyzed a decision tree model on the COVID-19 dataset, considering 42 selected features, including symptoms, using 10-fold cross-validation, and achieved an accuracy of 90.63% (6). Another study compared two decision tree models created by different classifier algorithms using Weka (7).

The objective of this study is to perform a comparative analysis of two decision trees on two distinct COVID-19 symptom datasets. By evaluating the performance and feature importance of these algorithms, we aim to gain insights into their predictive capabilities and their ability to identify key factors driving infection patterns.

## Methods

To assess the performance of decision trees, we employed two COVID-19 infection datasets that represent different infection scenarios. These datasets were thoughtfully chosen to encompass a wide range of symptoms and capture various factors influencing infection outcomes during the year 2020-2022.

### Datasets

#### Dataset acquisition and preprocessing

The first open data from the COVID Symptom Study - Sweden was downloaded from https://github.com/csss-resultat/covidsymptom in the R package: covidsymptom. For the second dataset, we utilized individual cases from a more recent timeframe, specifically around February-March 2022, sourced from the same data used in the previous study on machine learning for diagnosing COVID-19 infection (5). However, it is important to note that in our study, we maintained all the original classes present in the data, without excluding or modifying any of them.

The Israeli public open data on COVID-19 symptom was acquired from https://data.gov.il/dataset/covid-19. In the case of the Israeli dataset, we preserved the gender variable in its original format with three levels: male, female, and other. Subsequently, we examined the class distribution of the dataset, assessing whether it was balanced or imbalanced, by generating a bar plot. This analysis was conducted following a guide provided by finnstats (8). During the preprocessing stage, we performed data cleaning and removed unsuitable entries such as those with missing values or NAs. To create a more balanced dataset, we applied the Synthetic Minority Over-sampling Technique (SMOTE). This technique generates synthetic samples to address class imbalance and improve the representation of minority classes.

### Learning curve determination

We utilized the learning curve function in the caret package to analyze both the original dataset and the SMOTE-balanced dataset. Subsequently, we identified the dataset that produced the most favorable learning curve and used it to build the decision trees.

### Classifying and Hyperparameter tuning

The datasets were divided into a 75% training set and a 25% test set. We employed the train() function from the caret package to construct and fine-tune a model. Additionally, we employed a k-fold cross-validation approach with k=10 (10-fold cross-validation) to assess the model’s performance. To optimize the .cp parameter in the rpart model, we set the TuneLength to 20. For the random forest package, we utilized the rf method, while for the xgboost package, we employed the xgbTree method with default values for all seven hyperparameters.

When working with the SMOTE-balanced Israeli dataset, we exclusively utilized extreme gradient boosting to develop the final decision tree model. However, with the original Israeli dataset, we employed three classifiers (rpart with .cp parameter, random forest with .mtry parameter, and xgbTree model with seven parameters) to enhance the model’s performance.

### Evaluating the model

We evaluated the performance of all decision tree models by generating confusion matrices using the ConfusionMatrix() function. We compared their performance based on these metrics. Furthermore, we employed the ROCR package to generate receiver operating characteristic curves (ROC curves) in order to assess the performance of the models.

### Determination of Feature importance

We performed an extensive analysis of feature importance to identify the crucial variables that drive the predictions of the two decision trees applied to the Israeli and Swedish datasets. This analysis was conducted using the varImp() function from the caret package. By quantifying the contribution of each feature in the decision-making process, we gained valuable insights into the factors that play a significant role in shaping infection patterns. Understanding the importance of these features enhances our comprehension of the underlying mechanisms and facilitates the development of targeted interventions and preventive strategies.

## Results

The Swedish COVID-19 symptom dataset comprises 8,690 observations gathered between April 29th, 2020, and July 10th, 2022. All of these observations were included in the study due to their high quality and cleanliness. On the other hand, the Israeli COVID-19 symptom dataset initially consisted of 100,231 observations collected between February 17th, 2022, and March 3rd, 2022. However, after removing observations with missing age data, a total of 85,903 observations were utilized in the analysis.

We employed bar plotting to analyze the class distribution in the datasets. The class distribution of the Israeli dataset, depicted in Figure 1, exhibited a skew towards the negative class. Similarly, the class distribution in the Swedish dataset was not equal or close to balanced. To address this issue, we generated a new dataset with more balanced classes by applying SMOTE before modeling. Given that the imbalance appeared more severe in the first dataset compared to the second dataset, we utilized higher values for the perc.over and perc.under parameters during the SMOTE process.

**Figure 1.**
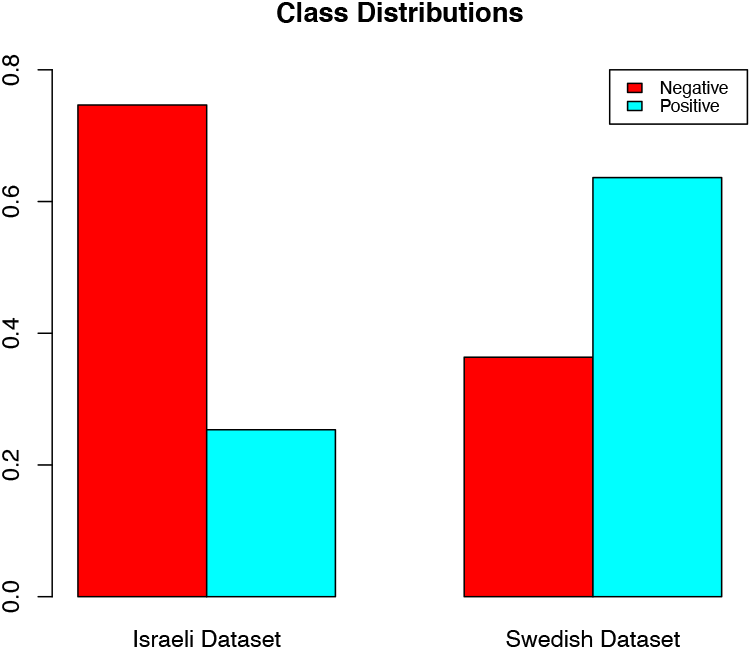
Class Distributions

Following the balancing of the datasets using various parameter sets, learning curves were generated for each of these datasets. To facilitate a comparison between the original dataset and two SMOTE-balanced datasets, we presented them side by side in Figure 2 (Swedish dataset) and Figure 3 (Israeli dataset).

**Figure 2.**
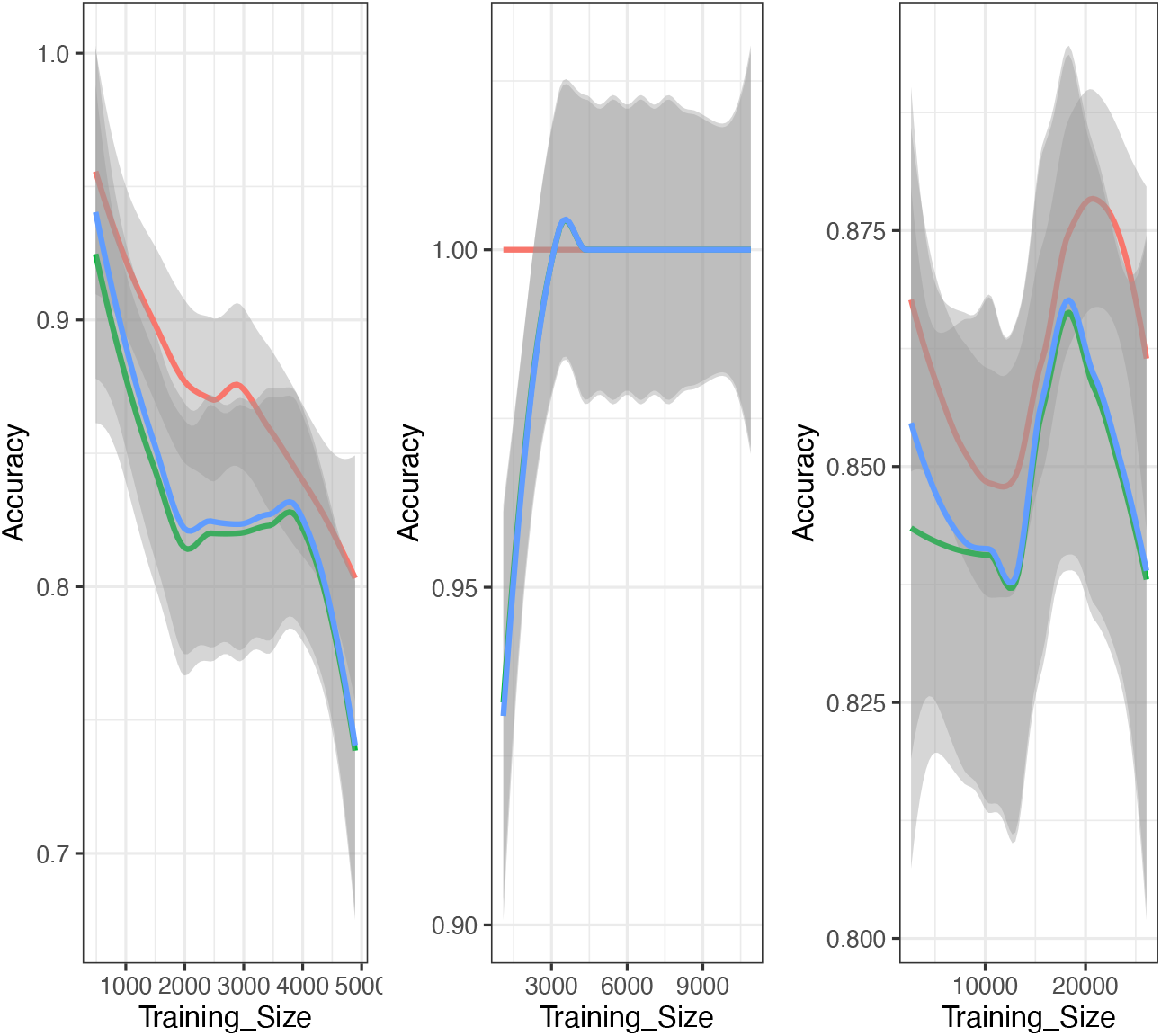
Learning curves depicting the performance of three Swedish datasets: the original actual dataset (Left), augmented with synthetic data; perc.over = 250, perc.under = 80 (Middle), and augmented with synthetic data; perc.over = 400, perc.under = 100 (Right). Legend lines indicate Resampling (Red), Testing (Green), and Training (Blue) iterations.

**Figure 3.**
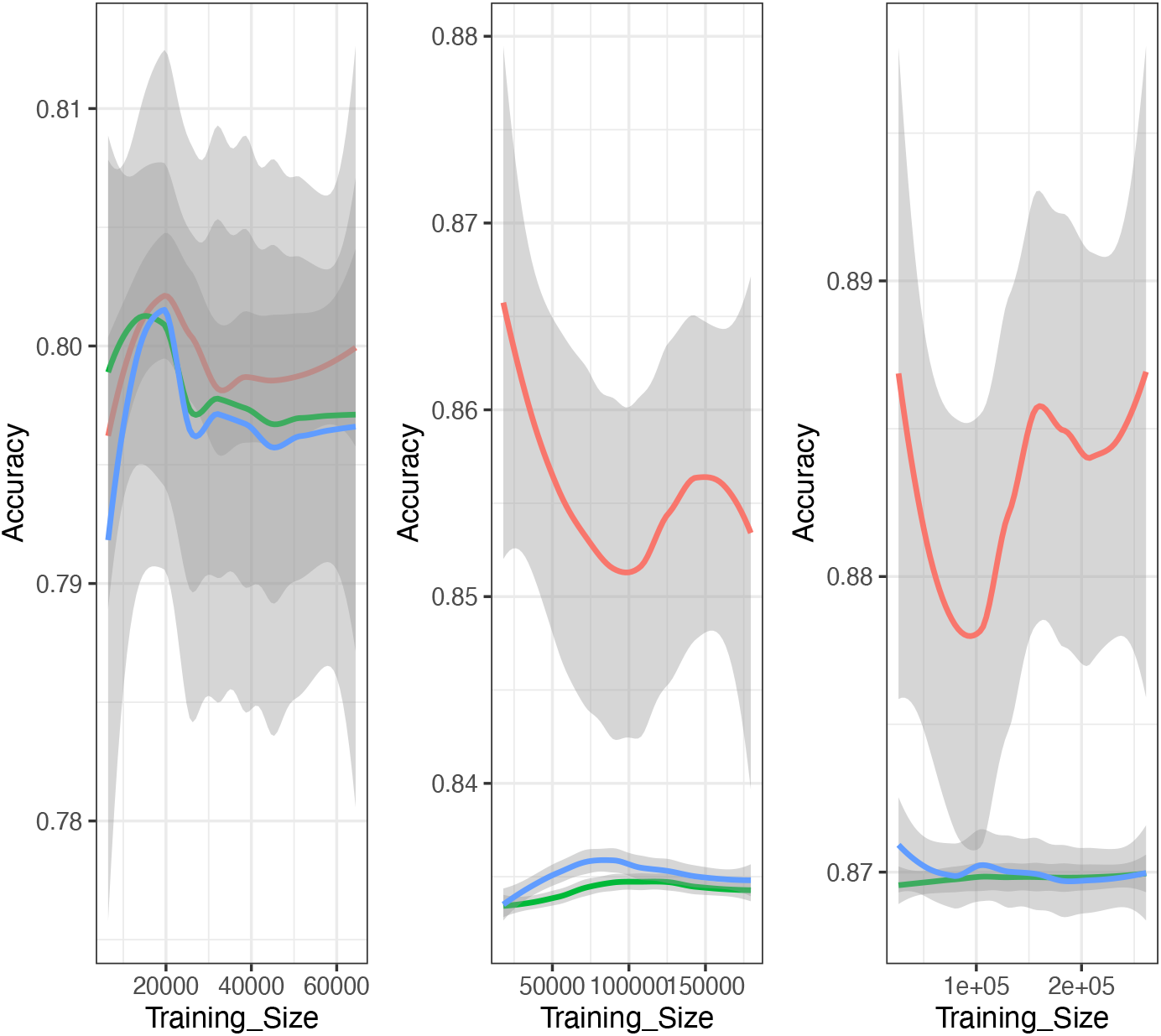
Learning curves illustrating the performance of three Israeli datasets: the original actual dataset (Left), augmented with synthetic data; perc.over = 400, perc.under = 100 (Middle), and augmented with synthetic data; perc.over = 500, perc.under = 200 (Right). Legend lines represent Resampling (Red), Testing (Green), and Training (Blue) iterations.

The models were created using balanced datasets that exhibited the best learning curves. The decision tree for the balanced Swedish dataset was built using the rpart method and 10-fold cross-validation (refer to Figure 4), whereas the balanced Israeli dataset was used to construct the model using extreme gradient boosting (see Figure 5).

**Figure 4.**
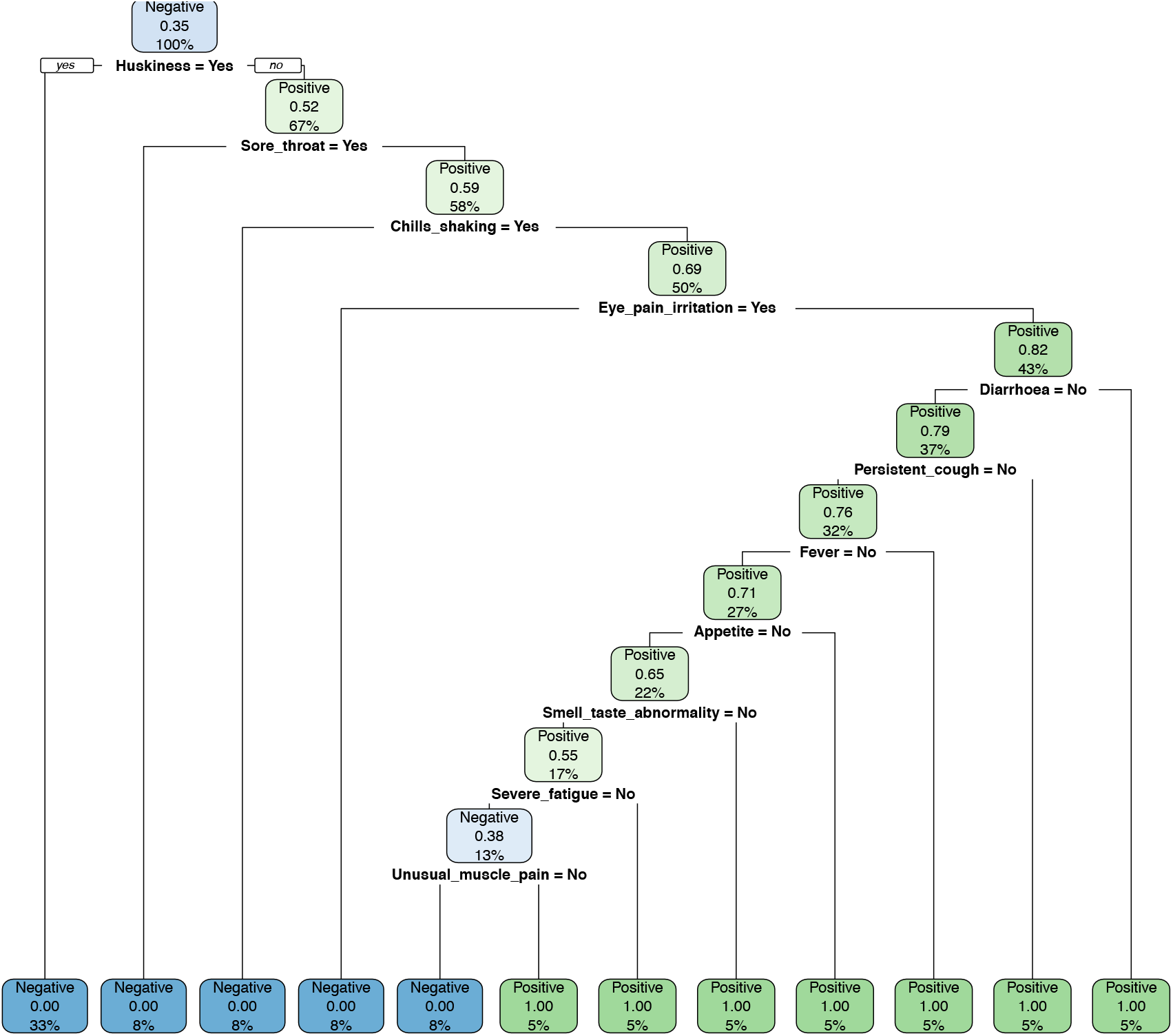
Decision Tree built from 250:80 combination-balanced Swedish dataset.

**Figure 5.**
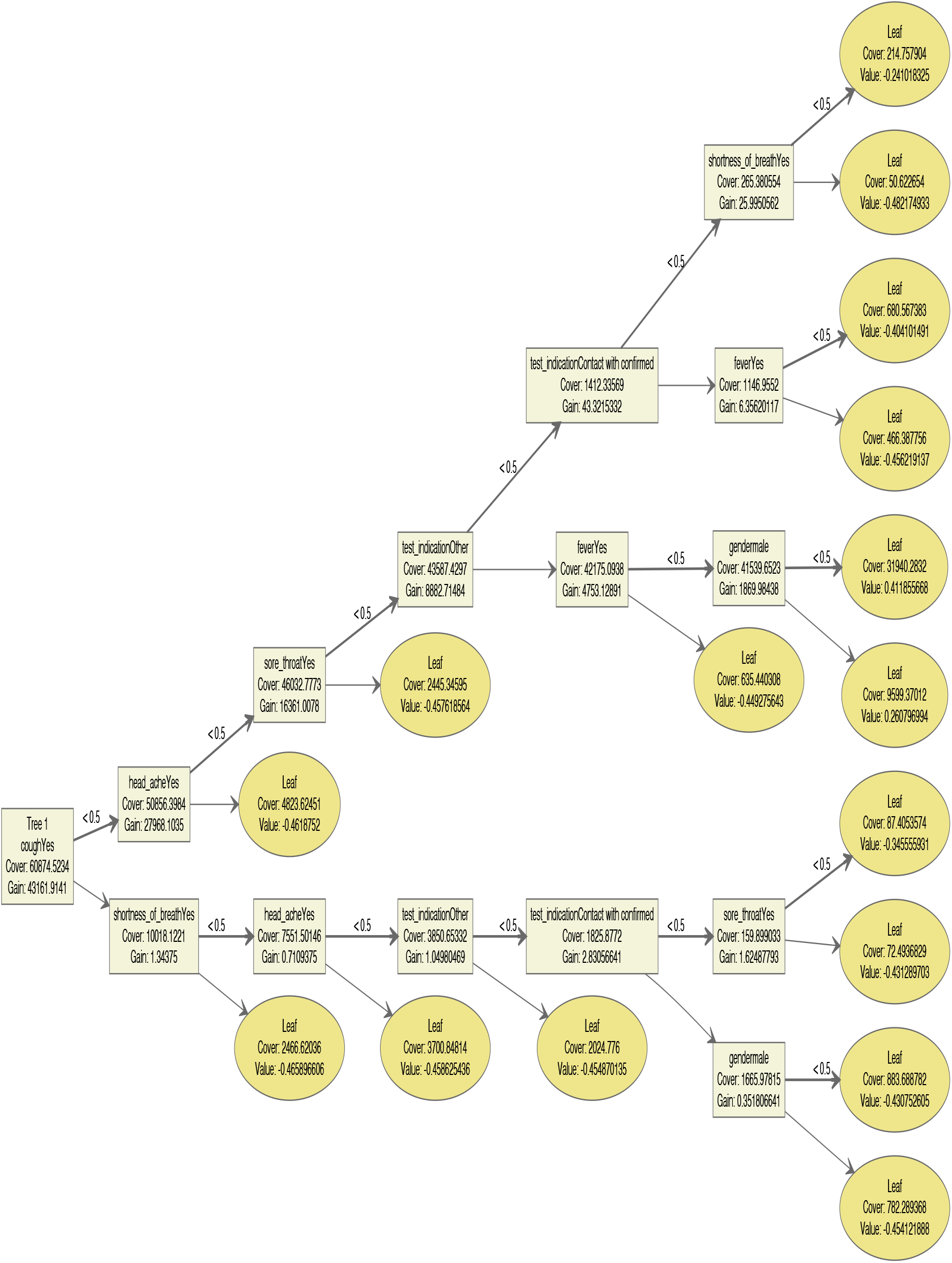
Decision tree created using the xgbTree method on the 500:200 combination-balanced Israeli dataset.

We then compared the performance of decision trees between original dataset and balanced dataset for both Swedish and Israeli datasets in Table 1.

**Table 1.** Performance metrics using Confusion matrix() function.

| Parameters | Models |  |  |  |
| --- | --- | --- | --- | --- |
|  | Swedish model<br>(Original) | Swedish model<br>(balanced<br>250:80) | Israeli model<br>(Original,<br>parameter tuned<br>with xgb Tree<br>method) | Israeli model<br>(balanced<br>500:200 with<br>xgb Tree<br>method) |
| Accuracy | 1 | 1 | 0.8125 | 0.9433 |
| Kappa | 1 | 1 | 0.3548 | 0.8758 |
| Sensitivity | 1.0000 | 1.0000 | 0.27871 | 0.8579 |
| Specificity | 1.0000 | 1.0000 | 0.99376 | 0.9946 |
| Precision | 1 | 1 | 0.9382 | 0.9896 |
| Recall | 1 | 1 | 0.2769 | 0.8582 |
| F1-score | 1 | 1 | 0.4276 | 0.9192 |

Due to the suboptimal performance of the Israeli original dataset, we sought to enhance the model’s performance by tuning its parameters using three different classifier methods. Table 2 provides a summary of the performance of these attempts, comparing them to the results obtained without any tuning.

**Table 2.** Hyperparameter tuning on the Israeli original model. The corresponding performance metrics were obtained using the confusionMatrix() function.

| Hyperparameters | Without tuning | Complexity parameter | mtry | Seven hyper-parameters in xgbTree method |
| --- | --- | --- | --- | --- |
| Accuracy (%) | 79.53 | 81.23 | 81.23 | 81.25 |
| Kappa | 0.2727 | 0.3539 | 0.3538 | 0.3548 |
| Sensitivity (%) | 20.83 | 27.78 | 27.76 | 27.87 |
| Specificity (%) | 99.45 | 99.38 | 99.39 | 99.38 |

We also assessed the performance of various models on two datasets: the original Israeli dataset and the Swedish dataset, using ROC curves (Figures not shown). These curves visually represent how well the models can distinguish between different classes. For the Israeli dataset, we considered four models: one without parameter tuning, one with a tuned .cp parameter, one with tuned parameters using the xgbTree method, and one with a tuned .mtry parameter using the rf method.

Analyzing the Israeli dataset, we observed that the models with .cp and xgbTree exhibited similar performance, as their ROC curves overlapped. This suggests that these models performed comparably in terms of classification ability. However, the models without tuning and .mtry had ROC curves closer to the diagonal line, indicating less effective performance compared to the other models.

Shifting our attention to the Swedish dataset, the ROC curve (Figure not shown) provided an overall assessment of the model’s performance. Notably, the curve was positioned closer to the top-left corner. This positioning indicates that the model achieved better overall performance on the Swedish dataset compared to the Israeli dataset.

To highlight the symptoms that are important in each dataset, we used varImp() function in the caret in determination of feature importance (Table 3).

**Table 3.**
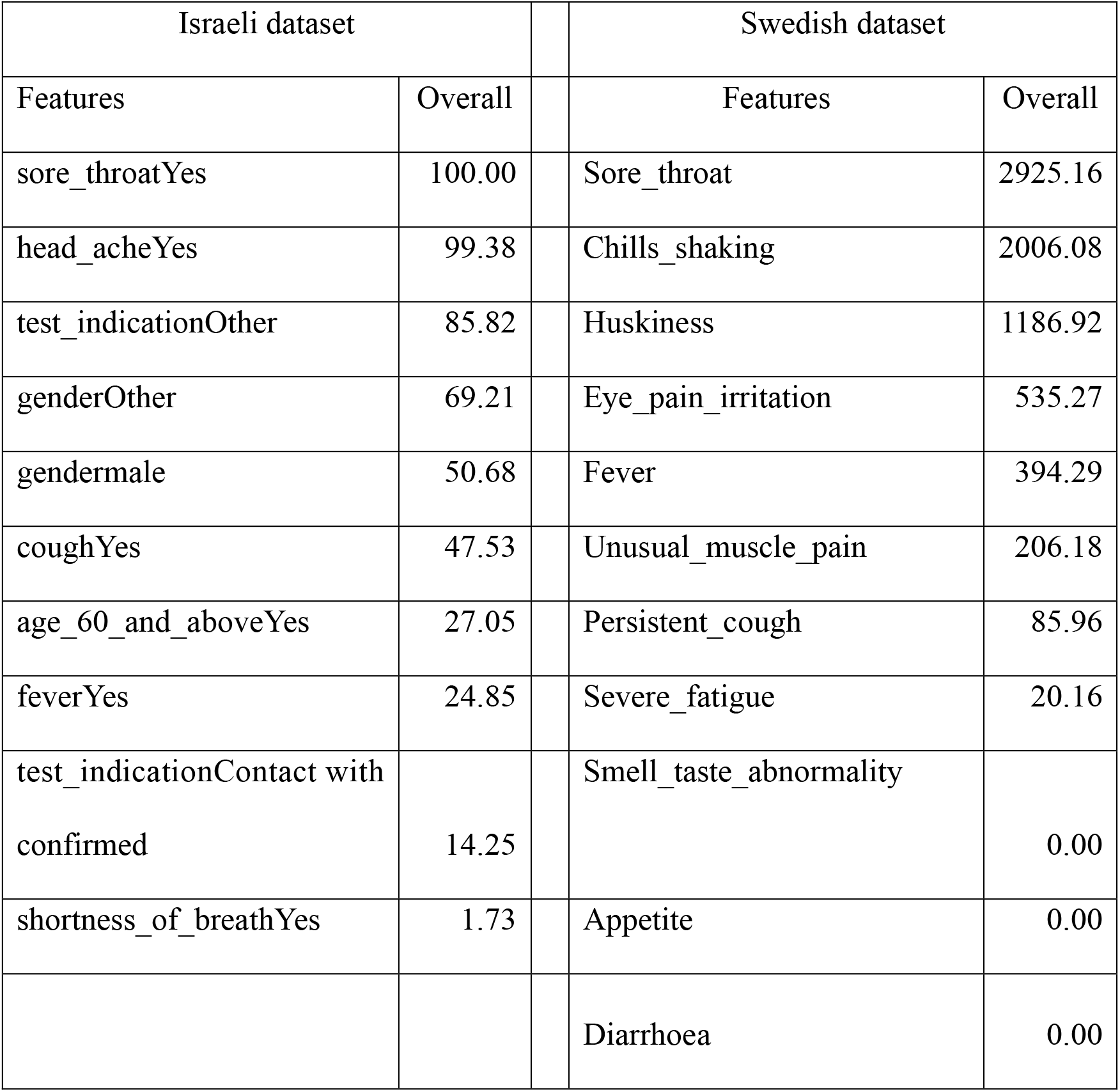
The important features in Swedish and Israeli datasets.

## Discussions and Conclusions

Decision tree classifiers operate by dividing the dataset into rectangular regions and constructing a basic model within each region. For our study, we employed the CART (Classification and Regression Tree) algorithm. The CART algorithm is widely recognized and utilized, and even the newer development of its major competitor (C5.0) has become quite similar to CART (4).

According to Swedish data, the class distribution ratio was 0.35:0.65. Therefore, we decided to balance the data using the perc.over range of 200-300 and perc.under range of 50-100. On the other hand, Israeli data showed a ratio of 1:3, so we opted for a perc.over range of 300-500 and perc.under range of 100-200. For this study, we utilized the SMOTE() function in DMwR. Initially, we set perc.over to 400 and perc.under to 150 for the first dataset (Israeli) and 250 and 80 for the second dataset (Swedish). These values served as starting points, and we explored various other combinations during the analysis.

By adopting the resampling line (Red) as a benchmark for interpreting the learning curves, we successfully determined the dataset suitable for modeling. The learning curves for the Swedish data (Figure 2) revealed that the dataset balanced with a 250:80 combination presented the most optimal balance for modeling purposes. Conversely, the Israeli dataset demonstrated that the best dataset was achieved through balancing with a 500:200 combination, as depicted in Figure 3.

In our meticulous examination of the model’s performance (Table 1), we made an intriguing observation. The Swedish dataset exhibited the most remarkable performance, even without the implementation of SMOTE transformation. However, when the Israeli dataset was balanced using a 500:200 combination, it demonstrated a significantly superior performance. Through meticulous hyperparameter tuning using the xgbTree method on the original Israeli dataset, we achieved a remarkable accuracy of 81.25%, a kappa value of 0.3548, a sensitivity of 27.87, and a specificity of 99.38.

Our study examined the performance of different models on the Israeli and Swedish datasets. The analysis of ROC curves revealed that the models with .cp and xgbTree had similar performance on the Israeli dataset, while the models without tuning and .mtry performed less effectively. Furthermore, the model demonstrated better performance on the Swedish dataset, as evident from the positioning of the ROC curve.

The introduction of SMOTE to balance the Israeli dataset resulted in a remarkable enhancement in multiple evaluation metrics, including accuracy, kappa value, specificity, sensitivity, precision, recall, and F1 score, when utilizing the xgbTree method. On the other hand, the Swedish COVID-19-symptom dataset, consisting of 8,690 observations (5,530 positive, 3,160 negative), demonstrated a high accuracy model even without hyperparameter tuning. However, after balancing the dataset, the learning curve indicated a significant improvement in the model’s performance. Given that the rpart method yielded the best performance on the Swedish dataset, we refrained from utilizing other classifier models.

Our research findings indicate that sore throat emerged as the most prominent symptom in both datasets. This observation is corroborated by a report that highlights sore throat as one of the four commonly experienced symptoms in individuals infected with the new variant (9). It is noteworthy that the global systematic review and meta-analysis conducted up to April 2020, comparing clinical findings of confirmed COVID-19 with influenza type A and B, did not identify sore throat as a significant symptom (10). However, it is crucial to acknowledge that the presentation of symptoms can vary depending on the specific strain of the virus. Furthermore, the Swedish dataset reported a higher prevalence of additional symptoms.

In conclusion, the Swedish and Israeli COVID-19 symptom datasets exhibit differences in terms of the number and types of variables, collection timing, class distribution, and number of observations. The model trained on the Swedish dataset achieved full accuracy, specificity, and sensitivity without requiring tuning. On the other hand, the model built on the actual Israeli dataset demonstrated high accuracy and specificity but low sensitivity and kappa values, which improved when utilizing the extreme gradient boosting method with default hyperparameter values. When using the SMOTE-balanced Israeli dataset, the model produced by the xgbTree method exhibited improved overall performance, including sensitivity.

Generally, the Swedish model displayed simplicity in terms of the number of nodes and depth, whereas the model developed on the SMOTE-balanced Israeli dataset exhibited greater complexity. The feature that was found to be important in both models was sore throat, which is one of the four common symptoms observed in individuals infected with new variants of the virus. Our study revealed differences between the models trained on the two symptom datasets and identified a crucial symptom for COVID-19. This finding enhances our understanding of COVID-19 in different geographical regions and its symptom presentation, providing valuable insights for future research and response strategies.

## Data Availability

Datasets have been available on the internet. The analyzed data can be obtained upon request.

https://github.com/csss-resultat/covidsymptom

https://data.gov.il/dataset/covid-19

## Ethics declarations

The datasets used in this study do not contain any private or sensitive data and are publicly accessible through the internet. The Israeli dataset was obtained from a source that had previously been used in another study, which did not require Internal Review Board (IRB) approval. Consequently, this current study also does not require IRB approval. The guidelines outlined in Luo and colleagues (11) were followed in preparing this manuscript.

## Author contributions

The authorship for this study adheres to the recommendations outlined by the International Committee of Medical Journal Editors (ICMJE) as stated in their guidelines (https://www.icmje.org/recommendations/browse/roles-and-responsibilities/defining-the-role-of-authors-and-contributors.html). P.W. and P.W.M. were involved in the study design and dedicated time to discussing the results and drawing conclusions. They thoroughly reviewed all drafts and provided valuable suggestions for improvement. Furthermore, they have given their approval for the final version of the manuscript, affirming its accuracy and integrity.

S.S. played a significant role in various stages of the study, including data acquisition, analysis, and interpretation. They were responsible for writing each draft and determining the content to be included in the final version for submission. To ensure the accuracy, relevance, and integrity of the study, every draft underwent rigorous discussions with the research team, resulting in substantial improvements.

## Acknowledgements

We would like to express our gratitude to the Israeli and Swedish governments for providing the public datasets utilized in this study.

## Conflicts of interest

All authors declare none.

### Box 1: Study contributions

Our study compared decision trees using two COVID-19 symptom datasets: Swedish and Israeli. Surprisingly, the decision tree trained on the Swedish dataset performed better even without SMOTE-balancing and hyperparameter tuning. In contrast, the decision tree trained on the Israeli dataset, with SMOTE transformation, showed significantly improved performance. Both models identified sore throat as a prominent symptom in the new virus variant, along with three others, as important features for detecting COVID-19 cases.

Implementing effective decision tree models in health services can predict COVID-19 infection likelihood. Public health agencies can use these models to enhance surveillance measures and incorporate the prominent symptom of sore throat as a crucial indicator. By integrating this information into their protocols, agencies can better monitor and respond to potential outbreaks.

To provide future insights based on our findings, it is essential to test the decision tree model using newer symptom datasets. This evaluation will enable us to assess its performance and determine if the identified important feature driving the model remains consistent. Conducting such tests will enhance the robustness of the model, making it more suitable for implementation in public health services. By continuously updating the dataset and evaluating the model’s performance, we can ensure its effectiveness in adapting to evolving situations and capturing any changes in the prominent symptoms associated with the virus. This iterative process will contribute to the reliability and relevance of the decision tree model when deployed in public health services.

